# Early warning system using primary healthcare data in the post-COVID-19-pandemic era: Brazil nationwide case-study

**DOI:** 10.1101/2023.11.24.23299005

**Authors:** Thiago Cerqueira-Silva, Juliane F. Oliveira, Vinicius de Araújo Oliveira, Pilar Tavares Veras Florentino, Alberto Sironi, Gerson O. Penna, Pablo Ivan Pereira Ramos, Viviane Sampaio Boaventura, Manoel Barral-Netto, Izabel Marcilio

## Abstract

**Background:** Syndromic surveillance utilising primary health care (PHC) data is a valuable tool for early outbreak detection, as demonstrated in the potential to identify COVID-19 outbreaks. However, the potential of such an early warning system in the post-COVID-19 era remains largely unexplored.

**Methods:** We analysed PHC encounter counts due to respiratory complaints registered in the Brazilian database of the Universal Health System between January and July 2023. We applied EARS (variation C1-C2-C3) and EVI to estimate the weekly thresholds. An alarm was determined when the number of encounters exceeded the week-specific threshold. We used data on hospitalisation due to respiratory disease to classify weeks in which the number of cases surpassed predetermined thresholds as anomalies. We compared EARS and EVI’s efficacy in anticipating anomalies.

**Findings:** A total of 119 anomalies were identified across 116 immediate regions during the study period. The EARS-C2 presented the highest early alarm rate, with 81/119 (68%) early alarms, and C1 the lowest, with 71 (60%) early alarms. The lowest true positivity was the EARS-C1 118/1354 (8.7%) and the highest EARS-C3 99/856 (11.6%).

**Conclusion:** Routinely collected PHC data can be successfully used to detect respiratory disease outbreaks in Brazil. Syndromic surveillance enhances timeliness in surveillance strategies, albeit with lower specificity. A combined approach with other strategies is essential to strengthen accuracy, offering a proactive and effective public health response against future outbreaks.

## Introduction

The COVID-19 pandemic has underscored the importance of timely and accurate surveillance systems in detecting and responding to emerging infectious diseases.^1,2^ Traditional surveillance methods, such as relying on laboratory-confirmed cases and hospital data, have timeliness, representativeness, and coverage limitations.^3,4^ Moreover, traditional surveillance systems do not prioritise early warning, and their usefulness for early detection of outbreaks has not been established.^5^

Syndromic surveillance systems were implemented to help provide situational awareness and inform patterns of illness distribution in the population.^6,7^ Syndromic surveillance relies on a set of pre-defined diagnostic symptoms, which are available before laboratory pathogen identification, therefore adding timeliness and sensitivity to the surveillance system. In this sense, integrating Primary Health Care (PHC) data into syndromic surveillance systems is regarded as a valuable source of information for early outbreak detection.^8,9^

The advancements in technology and the increasing availability of routinely collected health data highlight the importance of integrating digital health approaches to establish early warning systems for pandemic preparedness and response.^1,2^ The use of digital health in early warning enables the collection and analysis of diverse data streams. It represents a cost-effective solution by using data routinely gathered for healthcare and administrative purposes.

In the context of influenza-like illness (ILI), the importance of syndromic surveillance at the PHC level is particularly pronounced as the number of individuals with severe respiratory disease presenting to emergency rooms is expected to rise a few weeks after a marked increase in mild cases seeking PHC assistance. Brazil, with its vast population and comprehensive publicly funded healthcare system,^10^ provides an ideal setting to evaluate the potential of digital syndromic surveillance for anticipating ILI outbreaks. A previous study demonstrated the capabilities of using PHC data for the early detection of the COVID-19 first wave.^11^

This study aims to evaluate the potential of digital syndromic surveillance using PHC data for respiratory diseases to establish an early warning system in the post-COVID-19 pandemic era.

## Methods

### Study design

We evaluated the capabilities of an early warning system based on the weekly updated national PHC database using data on hospitalisations due to respiratory diseases as a gold standard. The study period went from October 2022 to July 2023. All analyses were aggregated by the geographic immediate region defined by the Brazilian Institute of Geography and Statistics (IBGE).^11^

### Data source

#### Primary Health Care Data

Brazil’s National Information System on Primary Health Care (SISAB) harbours data on all publicly funded PHC encounters in the country, coded by either the International Classification of Diseases (ICD-10) or the International Classification of Primary Care (ICPC-2). The PHC system covers at least 75% of the population in Brazil.^12^ Data for PHC encounters were extracted from the SISAB database and were obtained under the permission of the Ministry of Health (MoH). We used weekly counts of every PHC encounter due to ILI from October 2022 to July 2023. We included 50 ICD-10 and ICPC-2 codes corresponding to conditions possibly related to ILI. (Supplementary Table 1).

#### Hospital Information System

Brazil’s National Information System on Hospitalizations (SIH) comprises information on all publicly funded hospitalisations in Brazil, coded by the International Classification of Diseases (ICD-10). Data was extracted from January to July of 2023. We included 24 ICD-10 codes corresponding to respiratory conditions (Supplementary Table 2)

### Statistical Methods

We compared two methods to determine the threshold for an early warning in the PHC time series: the Early Aberration Reporting System (EARS, variations C1/C2/C3),^13^ and the Epidemic Volatility Index (EVI).^14^ The stark differences observed in the PHC time series between pre and post-COVID-19 years resulted in a non-stable baseline; thus an early warning system in the post-COVID-19 era required anomaly detection methods suitable for working with very short time series. The EARS method was developed by the United States Centers for Disease Control and Prevention to operate with short time series, requiring as little as 3 time points.^13^ The EVI was developed by Kostoulas et al. based on calculating the rolling standard deviation for a time series of confirmed COVID-19 cases and can also be employed with a short time series.^14^

We used an 8-week baseline for EARS and EVI, with a threshold alpha of 0.05 for the EARS and a threshold *c* of 0.1 for the EVI. Additionally, we conducted two sensitivity analyses: 1) changing the threshold alpha of 0.01 for EARS and c 0.2 for EVI to test the effect in specificity; and 2) using a 4-week baseline to test the effect in timeliness. An alarm in the PHC-based early warning system is established when the current number of encounters surpasses the week-specific threshold.

We defined an anomaly in the SIH time series by comparing the number of hospitalisations in the current week with the median number of hospitalisations per week in the study period. The thresholds for each immediate region were defined considering the median number of hospitalisations (Details on appendix section 1). Anomalies separated by one week alone were merged into one single event. Anomalies lasting for only one week were discarded, this criterion was applied to distinguish genuine events from random variation.

We evaluated the PHC-based early warning performance using three metrics derived from Nekorchuk et al. (2021)^15^: 1) Percent of events caught, here defined as the percent of anomalies in the SIH time series caught by the PHC-based early warning; 2) Percent of alarms associated with an anomaly (True positives): an alarm and anomaly were considered associated if the alarm was triggered any week during or up to three weeks prior to the anomaly; 3) Percent of timely alarms: defined as an alarm up to three weeks prior to the first week of an anomaly. We conducted the analysis stratified by population size of the immediate region categorised as small, medium, and large, as defined by the first and third quartiles. The weighted PHC coverage of each immediate region was calculated as the mean coverage of the municipalities comprising that region divided by the municipality’s population.

The statistical analysis was conducted using R software version 4.3.1 and the packages surveillance and EVI.

### Research ethics

The study is based on secondary, aggregated, non-identified data, and was approved by the Ethical Review Board of Oswaldo Cruz Foundation - Brasília Regional Office, CAAE 61444122.0.0000.0040.

## Results

Brazil has 510 immediate regions with populations ranging from 30,000 to 20 million inhabitants. The median population per region is 185,349 (interquartile range (IQR): 117,752 - 324,635). (Figure 1A) The weighted PHC coverage per immediate region ranged from 27% to 100%, with a median of 91% (IQR: 80 - 97). (Figure 1B)

**Figure 1:**
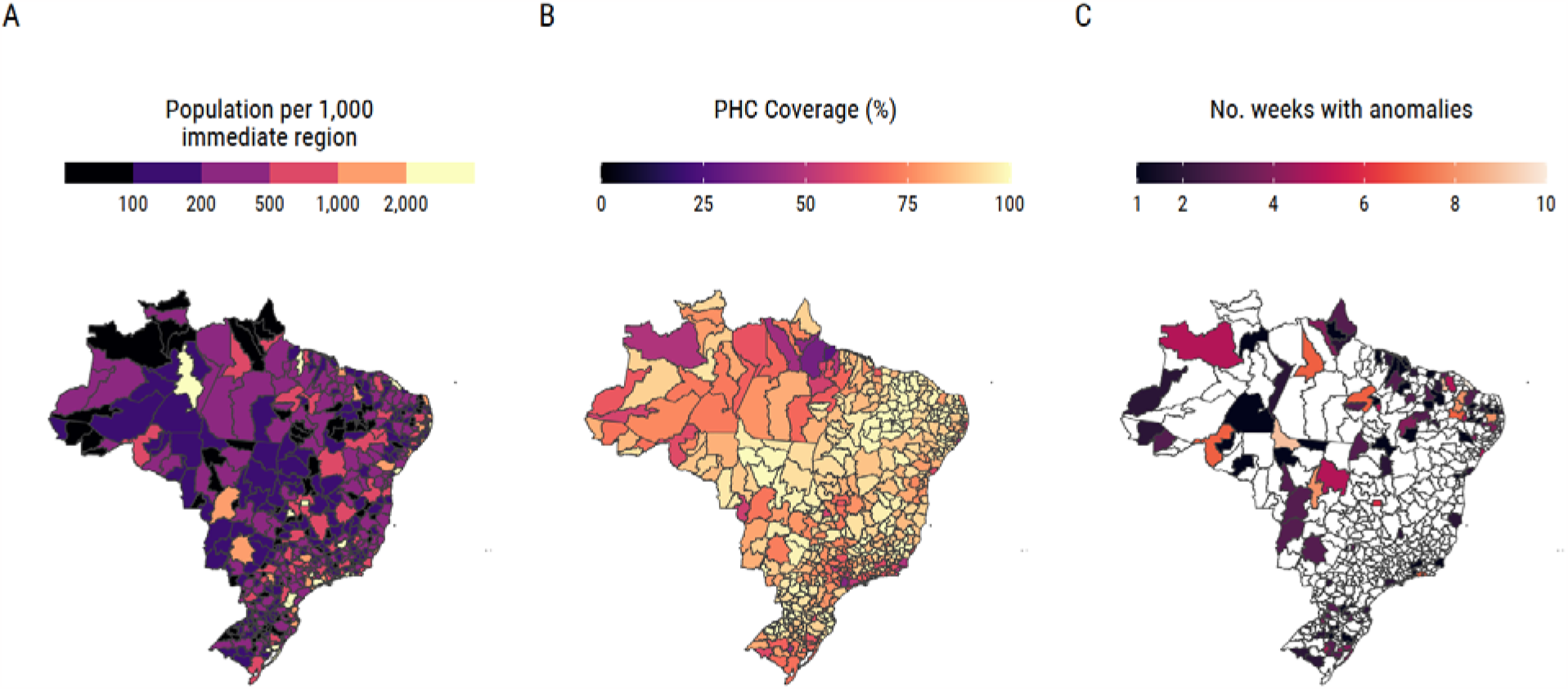
Brazil’s immediate regions. A) Population; B) Weighted PHC coverage; C) Number of weeks classified as anomaly between January to July of 2023.

The total number of PHC encounters steadily increased during the study period, peaking close to 10,000,000 encounters per week in June of 2023. The number of encounters due to ILI and hospitalisations due to respiratory causes also peaked around June 2023. (Figure 2 A-D)

**Figure 2:**
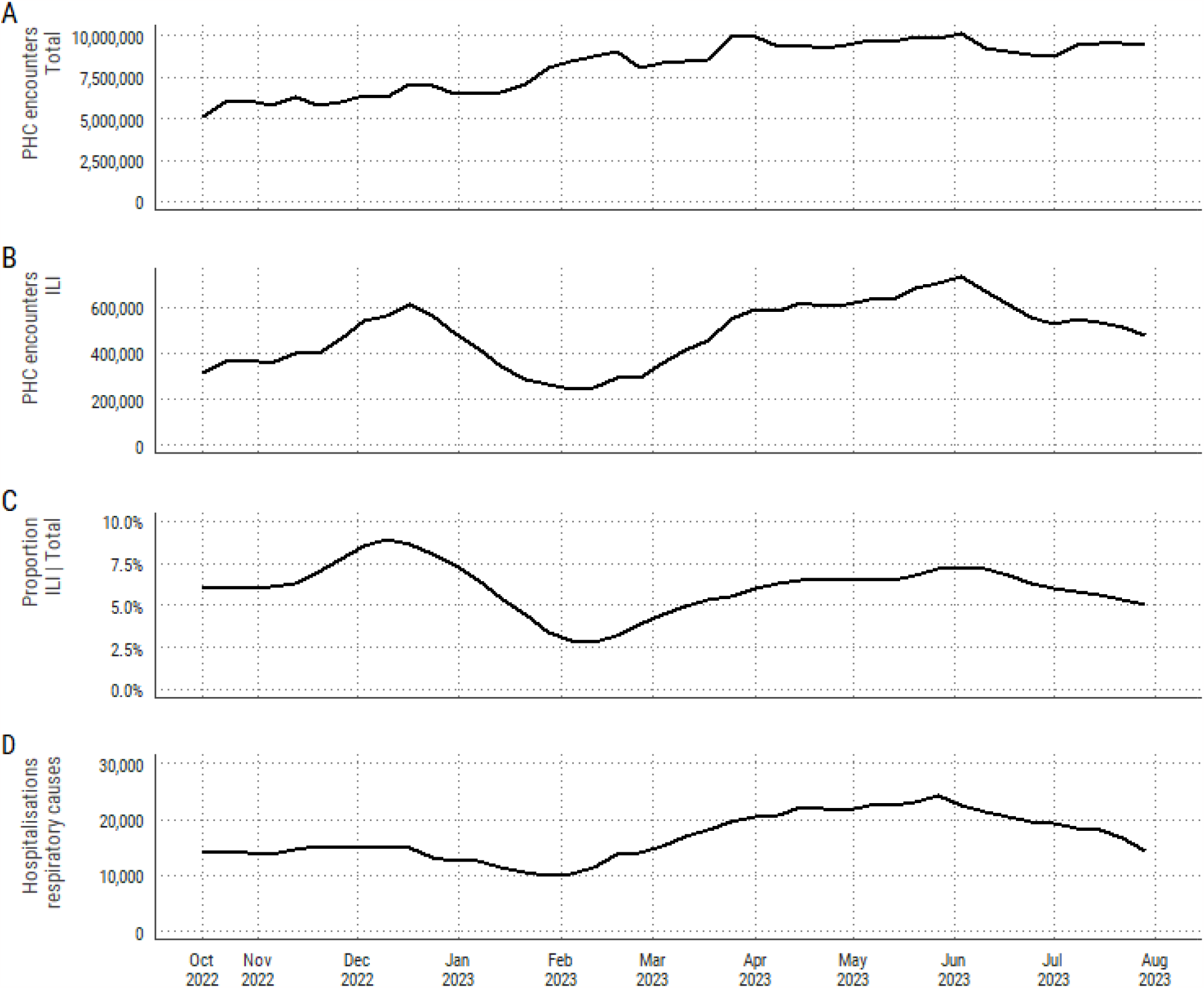
Encounters in the primary health care and hospitalisations due to acute respiratory causes (4-week moving average) between October 2022 and July 2023 in Brazil. A) Total encounters; B) Encounters related to influenza-like illness (ILI); C) Proportion of influenza-like illness encounters among total encounters; D) Number of hospitalisations due to acute respiratory causes

We identified 119 anomalies across 116 immediate regions in the SIH time series from January to July 2023, lasting from 2 to 11 weeks (Figure 1C and Supplementary Figure 1). The EARS-C2 presented the highest early alarm rate in the PHC time series, with 81/119 (68%) early alarms, and C1 the lowest, with 71 (60%) early alarms (Figure 3, Supplementary Table 3); 52 (44%) anomalies were early detected across all three variations of EARS and EVI, and 15 anomalies (13%) were not detected in any method. (Supplementary Table 4) Most missed anomalies in all methods last only two weeks (Supplementary Table 5). The true positivity was similar across all methods, ranging from 9% (EARS-C1) to 12% (EARS-C3). (Table 1)

**Figure 3:**
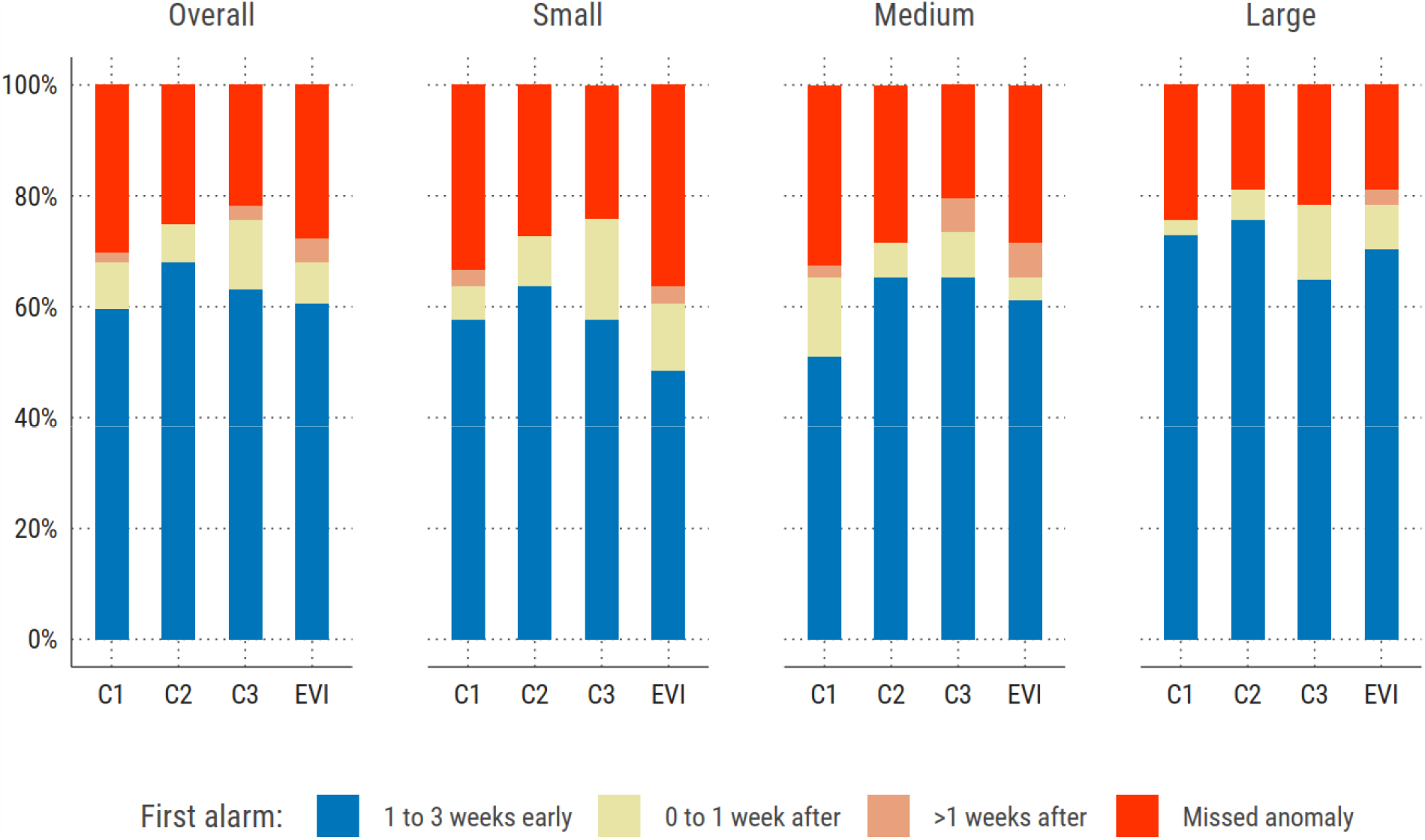
Performance of the methods, considering the time (in weeks) of the first alarm in the PHC series in relation to the first week with an anomaly in the SIH series, overall and stratified by population size.

**Table 1:**
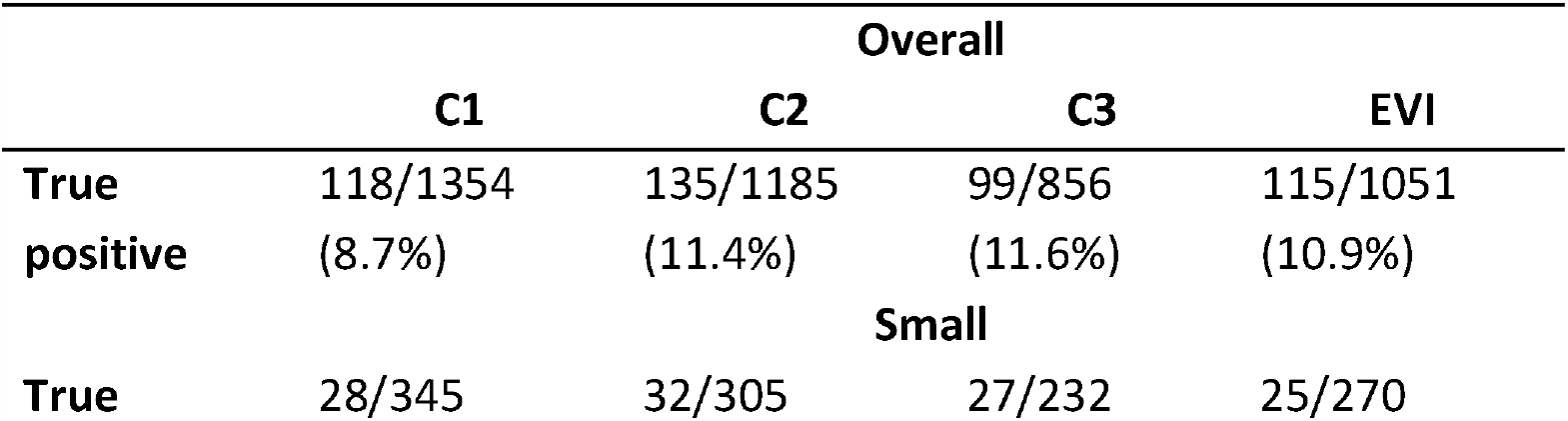
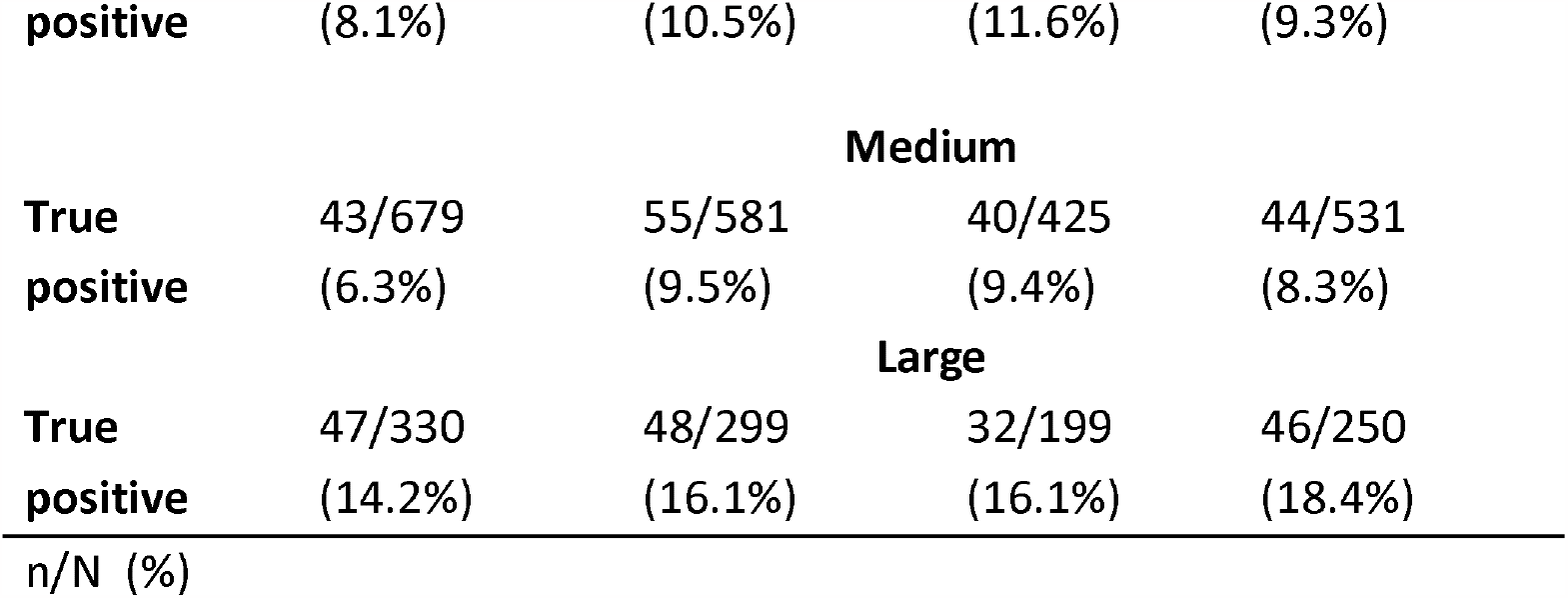
True positive rates per method. Overall and stratified by population size.

In the stratified analysis by population size, immediate regions with small populations had the lowest rate of early alarms, ranging from 48% to 64%, while regions with large populations had the highest rate, with values from 65 to 76%. (Figure 3) The true positivity rates were low across all strata, slightly better in large population regions, with values from 14 to 18%. (Table 1)

The sensitivity analysis using the threshold alpha of 0.01 for the EARS method and c 0.2 for the EVI did not increase the true positive rate. However, the early detection rate decreased for all methods, ranging from 36% (C1) to 56% (C2). (Supplementary Tables 7 and 8) The analysis using 4-week baseline improved the early detection rate for EARS (C2 and C3) and EVI, increasing from 68 to 72% (C2), 63 to 72% (C3), and 61 to 71% (EVI), and also improved caught rate in the same methods. The true positive rate decreased by 1 or 2% in all methods. (Supplementary Tables 7 and 8)

## Discussion

The findings of this study provide valuable insights into the longitudinal patterns of encounters due to respiratory causes in the PHC and the use of PHC data to develop an early warning system for respiratory disease outbreaks. We employed two methods for the early warning: EARS and EVI. Both methods showed capacity for early detection of respiratory disease outbreaks, with overall detection rate ranging from 60% to 68%. However, population size impacted the true positivity and early detection rate, with small population regions presenting the lowest number of true positives and early alarms.

The EARS and EVI methods offer flexibility for use in situations with limited historical data by relying on recent information for threshold setting. However, they exhibit a notable drawback: a decreased ability to accommodate seasonality, resulting in alarms often triggered during seasonal peaks. Methods able to adjust for seasonality, such as the improved Farrington method, tend to perform better in the true positive metric.^16^ The sustained high fluctuations of PHC encounters due to COVID-19 cases from 2020 to 2022, with misleading low numbers during lockdown periods, hinder the use of methods that require longer historical data as a baseline.^17,18^ Bédubourg and Le Stratt (2017)^16^ compared 21 early warning methods using simulated datasets and found that the probability of detection (POD) of an outbreak ranged from 43.3 to 84.4% and the false positive rate ranged from 0.7 to 59.9%. The EARS variations showed a POD ranging from 54.2 to 68.0% and a FPR of 6.9 to 8.5%. Similar to our findings, the C2 EARS variation presented the best performance metrics. The authors concluded that no single method presented outbreak detection performances sufficient enough to provide reliable monitoring for a large surveillance system.^16^ Using real surveillance data, Nekorchuk et al. (2021)^15^ compared three early warning methods for detecting malaria outbreaks. They found that the improved Farrington method showed the most effective results, as it could achieve the best trade-off on maximising both sensitivity (>70%) and specificity (>70%). Similar to our study, when analysing the three EARS variations, they found a high percentage of events caught (80 to 100%), with a moderate early detection rate (43 to 87%) and a low true positive rate (25 to 40%).^15^

In the context of an article on early warning systems, it is important to highlight the distinct advantages of integrating PHC data with conventional surveillance systems.^8,9^ This is particularly relevant in Brazil, where the granularity of PHC is exceptionally valuable. PHC extends its reach even to regions where more advanced healthcare facilities are lacking, reaching underserved rural and remote regions.^10^ The granularity of PHC plays a pivotal role in offering a timely window for detecting alarms through syndromic surveillance. It enables the early recognition of emerging health threats, even in areas with limited access to higher complexity healthcare infrastructure. This, in turn, allows for more rapid responses and proactive public health measures, ultimately enhancing the resilience of the healthcare system and safeguarding the well-being of vulnerable populations.^19^

The performance metrics shown here should be interpreted within the context of syndromic surveillance early alarms. It is generally accepted that syndromic surveillance provides high sensitivity but low specificity,^6^ and that the usefulness of early warning systems resides in indicating that an aberrant situation is occurring and should be investigated by health authorities.^20^ An early warning system complements, but does not replace, conventional surveillance strategies. The choice of which method to use depends on the data availability and which detection characteristics are most important in a given situation.^21^ Different parameterisations of detection methods will yield different performances, and, in general, a trade-off between the power of detection, false positive rates and early detection should be considered when choosing a particular method.^15,22,23^

Our study presents potential limitations. First, we relied on an algorithmic approach using reported hospitalisations due to acute respiratory causes to as a gold standard for defining outbreaks. Thus, surges of respiratory diseases that do not progress to severe disease would not be considered an outbreak in this study, and the signal detected in the PHC data would be considered a false positive. Second, we could not link individual PHC encounters and SIH data, preventing us from estimating the percentage of individuals who first sought primary health care before developing severe symptoms.

In conclusion, this study highlights the value of leveraging digital syndromic surveillance for the early detection of outbreaks. It offers valuable insights into utilising routinely collected PHC data for respiratory disease outbreak detection in Brazil. Working in this endeavour is crucial for enhancing surveillance accuracy and mitigating future outbreaks. This study contributes to the growing body of knowledge essential for addressing the complex challenges posed by infectious diseases, thus promoting a more proactive and effective public health response.

## Data Availability

Our agreement with the MoH for accessing the databases patently denies authorization of access to a third party. Any information for assessing the databases must be addressed to the Brazilian MoH at https://datasus.saude.gov.br/, and requests can be addressed to

