## Supplementary Material for "Early warning system using primary healthcare data in the post-COVID-19-pandemic era: Brazil nationwide case-study"

### Section 1: Additional Methods

Definition of an anomaly occurrence was defined as a function of the median (p50) of hospitalisations due to acute respiratory causes from January to July 2023. The rules for anomaly threshold are summarised in the table below. For handling random fluctuations in regions with a p50 less than 50 hospitalisations, the definition of an anomaly occurrence in a given week required a minimum of 10 hospitalisations in that week.

| Median (p50) of hospitalisations due to respiratory causes | Threshold |
| --- | --- |
| p50<50 | N > (2 * p50) & N>10 |
| 50 ≤ p50 <100 | N > (p50 + 0.5*p50) |
| 100 ≤ p50 <250 | N > (p50 + 0.4*p50) |
| 250 ≤ p50 <500 | N > (p50 + 0.3*p50) |
| 500 ≤ p50 <1000 | N > (p50 + 0.2*p50) |
| p50≥1000 | N > (p50 + 0.1*p50) |

### Supplementary Table 1: ICD-10 and ICPC-2 codes used to define conditions likely related to influenza-like illness.

| **Type** | **Code** | **Description** |
| --- | --- | --- |
| ICPC-2 | A03 | Fever |
| ICPC-2 | R01 | Pain respiratory system |
| ICPC-2 | R02 | Shortness of breath/dyspnoea |
| ICPC-2 | R03 | Wheezing |
| ICPC-2 | R04 | Breathing problem, other |
| ICPC-2 | R05 | Cough |
| ICPC-2 | R07 | Sneezing / nasal congestion |
| ICPC-2 | R08 | Nose symptom / complaint other |
| ICPC-2 | R21 | Sinus symptom / complaint |
| ICPC-2 | R23 | Voice symptom / complaint |
| ICPC-2 | R25 | Sputum / phlegm abnormal |
| ICPC-2 | R29 | Respiratory symptom/complaint other |
| ICPC-2 | R71 | Whooping cough |
| ICPC-2 | R74 | Upper respiratory infection acute |
| ICPC-2 | R75 | Sinusitis acute/chronic |
| ICPC-2 | R76 | Tonsillitis acute |
| ICPC-2 | R77 | Laryngitis/tracheitis acute |
| ICPC-2 | R78 | Acute bronchitis/bronchiolitis |
| ICPC-2 | R80 | Influenza |
| ICPC-2 | R81 | Pneumonia |
| ICPC-2 | R83 | Respiratory infection other |
| ICPC-2 | R99 | Respiratory disease other |
| ICD-10 | J00 | Acute nasopharyngitis |
| ICD-10 | J01 | Acute sinusitis |
| ICD-10 | J02 | Acute pharyngitis |
| ICD-10 | J03 | Acute tonsillitis |
| ICD-10 | J04 | Acute laryngitis and tracheitis |
| ICD-10 | J06 | Acute upper respiratory infections of multiple and unspecified sites |
| ICD-10 | J09 | Influenza due to identified zoonotic or pandemic influenza virus |
| ICD-10 | J10 | influenza due to identified seasonal influenza virus |
| ICD-10 | J11 | Influenza, virus not identified |
| ICD-10 | J12 | Viral pneumonia, not elsewhere classified |
| ICD-10 | J13 | Pneumonia due to Streptococcus pneumoniae |
| ICD-10 | J14 | Pneumonia due to Haemophilus influenzae |
| ICD-10 | J15 | Bacterial pneumonia, not elsewhere classified |
| ICD-10 | J16 | Pneumonia due to other infectious organisms, not elsewhere classified |
| ICD-10 | J17 | Pneumonia in diseases classified elsewhere |
| ICD-10 | J18 | Pneumonia, organism unspecified |
| ICD-10 | J20 | Acute bronchitis |
| ICD-10 | J21 | Acute bronchiolitis |
| ICD-10 | J22 | Unspecified acute lower respiratory infection |
| ICD-10 | J80 | Adult respiratory distress syndrome |
| ICD-10 | R05 | Cough |
| ICD-10 | R06 | Abnormalities of breathing |
| ICD-10 | R07 | Pain in throat and chest |
| ICD-10 | R43 | Disturbances of smell and taste |
| ICD-10 | R50 | Fever of other and unknown origin |
| ICD-10 | U07 | Emergency use of U07 |
| ICD-10 | B34 | Viral infection of unspecified site |
| ICD-10 | B97 | Viral agents as the cause of diseases classified to other chapters |

### Supplementary Table 2: ICD-10 codes used to define hospitalisations due to respiratory causes.

| **Type** | **Code** | **Description** |
| --- | --- | --- |
| ICD-10 | J00 | Acute nasopharyngitis |
| ICD-10 | J01 | Acute sinusitis |
| ICD-10 | J02 | Acute pharyngitis |
| ICD-10 | J03 | Acute tonsillitis |
| ICD-10 | J04 | Acute laryngitis and tracheitis |
| ICD-10 | J06 | Acute upper respiratory infections of multiple and unspecified sites |
| ICD-10 | J09 | Influenza due to identified zoonotic or pandemic influenza virus |
| ICD-10 | J10 | influenza due to identified seasonal influenza virus |
| ICD-10 | J11 | Influenza, virus not identified |
| ICD-10 | J12 | Viral pneumonia, not elsewhere classified |
| ICD-10 | J13 | Pneumonia due to Streptococcus pneumoniae |
| ICD-10 | J14 | Pneumonia due to Haemophilus influenzae |
| ICD-10 | J15 | Bacterial pneumonia, not elsewhere classified |
| ICD-10 | J16 | Pneumonia due to other infectious organisms, not elsewhere classified |
| ICD-10 | J17 | Pneumonia in diseases classified elsewhere |
| ICD-10 | J18 | Pneumonia, organism unspecified |
| ICD-10 | J20 | Acute bronchitis |
| ICD-10 | J21 | Acute bronchiolitis |
| ICD-10 | J22 | Unspecified acute lower respiratory infection |
| ICD-10 | J80 | Adult respiratory distress syndrome |
| ICD-10 | U07 | Emergency use of U07 |
| ICD-10 | B34 | Viral infection of unspecified site |
| ICD-10 | B97 | Viral agents as the cause of diseases classified to other chapters |
| ICD-10 | U04 | Severe acute respiratory syndrome |

### Supplementary Table 3: Performance of the methods, considering the time (in weeks) of the first alarm in the PHC series in relation to the first week with an anomaly in the SIH series, overall and stratified by population size.

|  | **Overall, N = 119** | | | | **Small, N = 33** | | | | **Medium, N = 49** | | | | **Large, N = 37** | | | |
| --- | --- | --- | --- | --- | --- | --- | --- | --- | --- | --- | --- | --- | --- | --- | --- | --- |
| **First Alarm** | **C1** | **C2** | **C3** | **EVI** | **C1** | **C2** | **C3** | **EVI** | **C1** | **C2** | **C3** | **EVI** | **C1** | **C2** | **C3** | **EVI** |
| 1 to 3 weeks early | 71 (59.7) | 81 (68.1) | 75 (63.0) | 72 (60.5) | 19 (57.6) | 21 (63.6) | 19 (57.6) | 16 (48.5) | 25 (51.0) | 32 (65.3) | 32 (65.3) | 30 (61.2) | 27 (73.0) | 28 (75.7) | 24 (64.9) | 26 (70.3) |
| 0 to 1 week after | 10 (8.4) | 8 (6.7) | 15 (12.6) | 9 (7.6) | 2 (6.1) | 3 (9.1) | 6 (18.2) | 4 (12.1) | 7 (14.3) | 3 (6.1) | 4 (8.2) | 2 (4.1) | 1 (2.7) | 2 (5.4) | 5 (13.5) | 3 (8.1) |
| >1 weeks after | 2 (1.7) | 0 (0.0) | 3 (2.5) | 5 (4.2) | 1 (3.0) | 0 (0.0) | 0 (0.0) | 1 (3.0) | 1 (2.0) | 0 (0.0) | 3 (6.1) | 3 (6.1) | 0 (0.0) | 0 (0.0) | 0 (0.0) | 1 (2.7) |
| Missed anomaly | 36 (30.3) | 30 (25.2) | 26 (21.8) | 33 (27.7) | 11 (33.3) | 9 (27.3) | 8 (24.2) | 12 (36.4) | 16 (32.7) | 14 (28.6) | 10 (20.4) | 14 (28.6) | 9 (24.3) | 7 (18.9) | 8 (21.6) | 7 (18.9) |
| n (%) | | | | |  |  |  |  |  |  |  |  |  |  |  |  |

### Supplementary Table 4: Number of anomalies that multiple methods detected early or missed.

| **Number of methods** | **Anomalies detected early** | **Anomalies missed** |
| --- | --- | --- |
| 0 | 22 (18.5) | 76 (63.9) |
| 1 | 14 (11.8) | 6 (5) |
| 2 | 16 (13.4) | 7 (5.9) |
| 3 | 15 (12.6) | 15 (12.6) |
| 4 | 52 (43.7) | 15 (12.6) |

**n(%)**

### Supplementary Table 5: Length of missed anomalies by method.

| **No. weeks** | **C1, N = 36** | **C2, N = 30** | **C3, N = 26** | **EVI, N = 33** |
| --- | --- | --- | --- | --- |
| **2** | 15 (42) | 14 (47) | 10 (38) | 15 (45) |
| **3** | 8 (22) | 5 (17) | 7 (27) | 9 (27) |
| **4** | 8 (22) | 7 (23) | 5 (19) | 6 (18) |
| **5** | 2 (5.6) | 2 (6.7) | 1 (3.8) | 2 (6.1) |
| **6** | 3 (8.3) | 2 (6.7) | 2 (7.7) | 1 (3.0) |
| **8** | 0 (0) | 0 (0) | 1 (3.8) | 0 (0) |
| **n (%)** | | | | |

### Supplementary Table 6: Performance of the methods in the two sensitivity analyses. N represent the total of anomalies.

|  | **Sensitivity – Change threshold values,**  **N = 119** | | | | **Sensitivity – Change baseline value,**  **N = 119** | | | |
| --- | --- | --- | --- | --- | --- | --- | --- | --- |
| **First Alarm** | **C1** | **C2** | **C3** | **EVI** | **C1** | **C2** | **C3** | **EVI** |
| 1 to 3 weeks early | 43 (36.1) | 67 (56.3) | 63 (52.9) | 56 (47.1) | 58 (48.7) | 86 (72.3) | 86 (72.3) | 85 (71.4) |
| 0 to 1 week after | 14 (11.8) | 10 (8.4) | 15 (12.6) | 11 (9.2) | 18 (15.1) | 6 (5.0) | 10 (8.4) | 8 (6.7) |
| >1 weeks after | 2 (1.7) | 2 (1.7) | 4 (3.4) | 4 (3.4) | 2 (1.7) | 2 (1.7) | 3 (2.5) | 8 (6.7) |
| Missed anomaly | 60 (50.4) | 40 (33.6) | 37 (31.1) | 48 (40.3) | 41 (34.5) | 25 (21.0) | 20 (16.8) | 18 (15.1) |
| n (%) | | | | |  |  |  |  |

### Supplementary Table 7: True positive rates per method of the two sensitivity analyses. N represent the total of alarm of each method.

|  | Sensitivity – Change threshold values | | | | Sensitivity – Change baseline value | | | |
| --- | --- | --- | --- | --- | --- | --- | --- | --- |
| **Characteristic** | **C1, N = 882** | **C2, N = 955** | **C3, N = 792** | **EVI, N = 808** | **C1, N = 1,806** | **C2, N = 1,500** | **C3, N = 1,064** | **EVI, N = 1,467** |
| True positive | 73 (8.3) | 105 (11.0) | 87 (11.0) | 82 (10.1) | 112 (6.2%) | 136 (9.1%) | 117 (11.0%) | 129 (8.8%) |

### Supplementary Figure 1: Length of anomalies stratified by population size

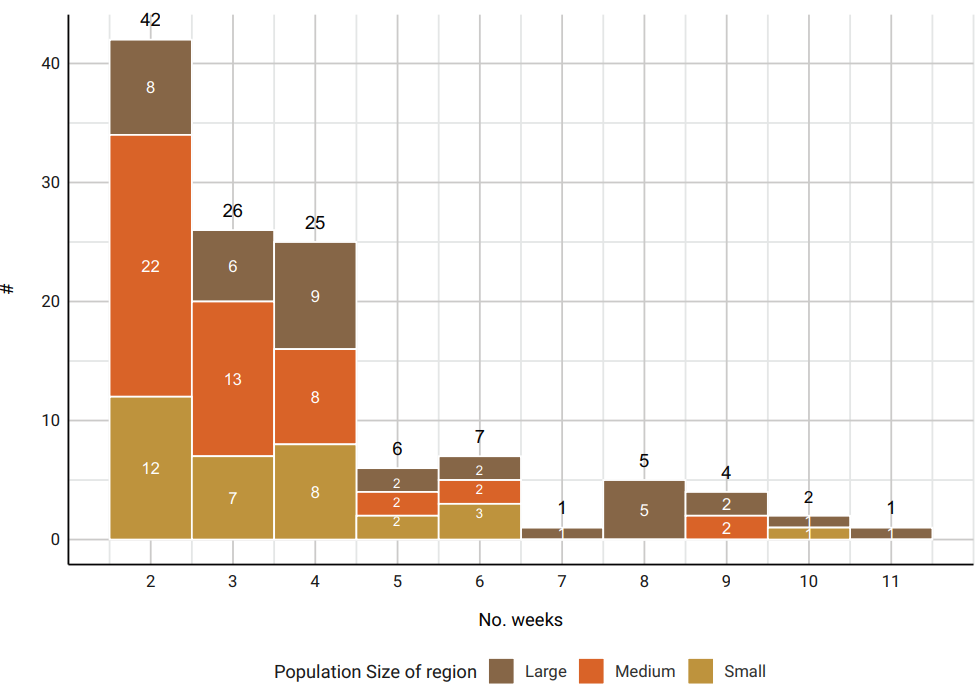
